# Estimation of offspring genetic risk scores using parental genotypes

**DOI:** 10.1101/2022.06.09.22276224

**Authors:** Adebayo Adesomo, Tsegaselassie Workalemahu, Matthew Givens, Mark Yandell, Aaron Quinlan, Martin Tristani-Firouzi, Sarah Heerboth, Robert Silver, Nathan Blue

**Affiliations:** University of Utah Health, Department of Obstetrics and Gynecology. 30 North 1900 East, 2B200. Salt Lake City, Utah 84132; Intermountain Healthcare. 5063 Cottonwood Street. Murray, Utah 84157; Department of Human Genetics, Utah Center for Genetic Discovery, University of Utah, Salt Lake City, USA; Department of Biomedical Informatics, University of Utah, Salt Lake City, USA; Division of Pediatric Cardiology, University of Utah, Salt Lake City, Utah, USA; Nora Eccles Harrison Cardiovascular Research and Training Institute, University of Utah, Salt Lake City, UT 84112, USA

## Abstract

**Objective:** Our objective was to determine whether genetic risk scores (GRSs) of offspring can be accurately estimated from parental DNA.

**Methods:** Whole genome sequencing data from a cohort of forty-seven multi-generation Utah families were used to extract single nucleotide polymorphism (SNP) data at genetic loci associated with the following traits: birth weight (BW), fasting plasma glucose (FPG), blood pressure (BP), body mass index (BMI), height, and type 2 diabetes (T2D). Offspring GRSs for each trait were estimated from parental single nucleotide polymorphism (SNP) data and compared to actual offspring GRSs. We also assessed offspring GRS estimation using only one parent’s DNA to simulate scenarios when only one genetic parent is available. The primary outcome was the percent error of parental-derived estimated GRS for each trait. An a priori threshold of 10% error was chosen for estimated GRSs to be considered accurate.

**Results:** Forty-three families with an average of 8.9 ± 1.8 offspring (N = 454 offspring) had parental and offspring SNP data available for GRS calculations. Mean percent errors for estimated offspring GRSs were less than 10% for all traits except for FPG (10.5% ± 8.1%). Percent errors were not significantly different when offspring GRSs were estimated using only one parent’s DNA whether the missing parent was a father or mother. Mean percent error of GRSs decreased exponentially with increasing SNPs per trait, with diminishing improvement in percent error above 500 SNPs.

**Conclusion:** Parental genetic risk scores can be used to accurately estimate genetic risk scores of offspring. This proof of concept supports further exploration of parental genetic risk scores as a tool for prenatal fetal genetic risk stratification.

**Statements:** What’s already known about this topic?

Genetic risk scoring is a tool to estimate the probability of development traits or conditions with complex, multifactorial inheritance.

What does this study add?

Offspring genetic risk scores can be accurately estimated using parental DNA. This proof of concept supports further exploration of parental genetic risk scores as a tool for prenatal fetal genetic risk stratification.

## Introduction

Prenatal assessment of the fetal genome is widely used as a means for fetal diagnosis and risk stratification^1^. An increasingly common approach to risk stratification for conditions and traits with multifactorial inheritance is the calculation of genetic risk scores (GRSs) from single nucleotide polymorphism (SNP) arrays^2,3^. GRSs are using SNPs an individual carries at loci known to be associated with a given trait, either as a simple sum of inherited SNPs (raw GRS) or with each SNP multiplied by coefficients expressing the strength of each SNP’s association with the trait (weighted GRS). Fetal GRS analysis has the potential to improve individualized assessment of multifactorial outcomes that are currently difficult to estimate well, such as fetal growth or perinatal morbidity^4^.

Currently, fetal GRS analysis is only possible through direct interrogation of fetal or placental DNA. This can be achieved through amniocenteses or chorionic villous sampling, invasive procedures which pose maternal and fetal risks^5^. Non-invasive methods can circumvent the risks associated with these procedures by isolating cell-free placental DNA in maternal plasma. However, platforms allowing for non-invasive genome-wide SNP arrays are not yet widely available for clinical application. Therefore, our primary objective was to determine whether offspring GRSs can be accurately estimated form parental DNA using a cohort with both parental and offspring DNA available. Our secondary objective was to characterize offspring GRS estimation in the absence of one parent.

## Methods

This is a secondary analysis of data from the Utah Centre D’etude du Polymorphism Humaine (CEPH) cohort. CEPH (Centre D’etude du Polymorphism Humaine) is an international research consortium established in 1984, which recruited 61 large families whose DNA served as the haplotype map of the CEU (Central Europe) population for the HapMap and 1000 genomes projects, including 47 Utah families (Utah Genome Reference Project)^6,7^. The Utah cohort is composed of 639 participants from 47 three-generation Utah pedigrees who contributed DNA and underwent phenotype assessment in the 1980s and 1990s. The full processes of family recruitment and selection, phenotype assessment, as well as the general history and scientific accomplishments of the project have been documented previously. Recruitment included members from three generations centered on the middle generation, such that participants included two partnered parents, all their available offspring, and both parents’ parents. A representative pedigree is included in Figure 1. When DNA was available from grandparents in a given family, individuals in the middle generation were analyzed as both parents and offspring. Peripheral blood samples were collected from all participants and underwent DNA extraction and whole genome sequencing (WGS) at 30X median depth of coverage using the Illumina HiSeqX (Illumina, San Diego, CA). The full description of the sequencing pipeline was previously described^8^. Single nucleotide polymorphisms (SNPs) were called using the Genome Analysis Toolkit (GATK) workflow^9^. For this study, genotypes of candidate SNPs were extracted using available WGS data were used to extract SNP data from all participants using pLink software. Candidate SNPs were genetic loci known to be associated with the following traits were evaluated: birth weight (BW), fasting plasma glucose (FPG), blood pressure (BP), BMI, height, and type 2 diabetes (T2D)^10^. These traits were selected because their coefficients were all derived from a common cohort using polygenic risk score methodology, and they are relevant for exploring implications of fetal growth with adult metabolic disease. GRSs were based on the following number of SNPs: height: 2,130; BP: 831; BMI: 628; T2D: 306; BW: 86; FPG: 22. All SNPs are autosomal except for two in the BW score, which are on the X chromosome.

**Figure 1.**
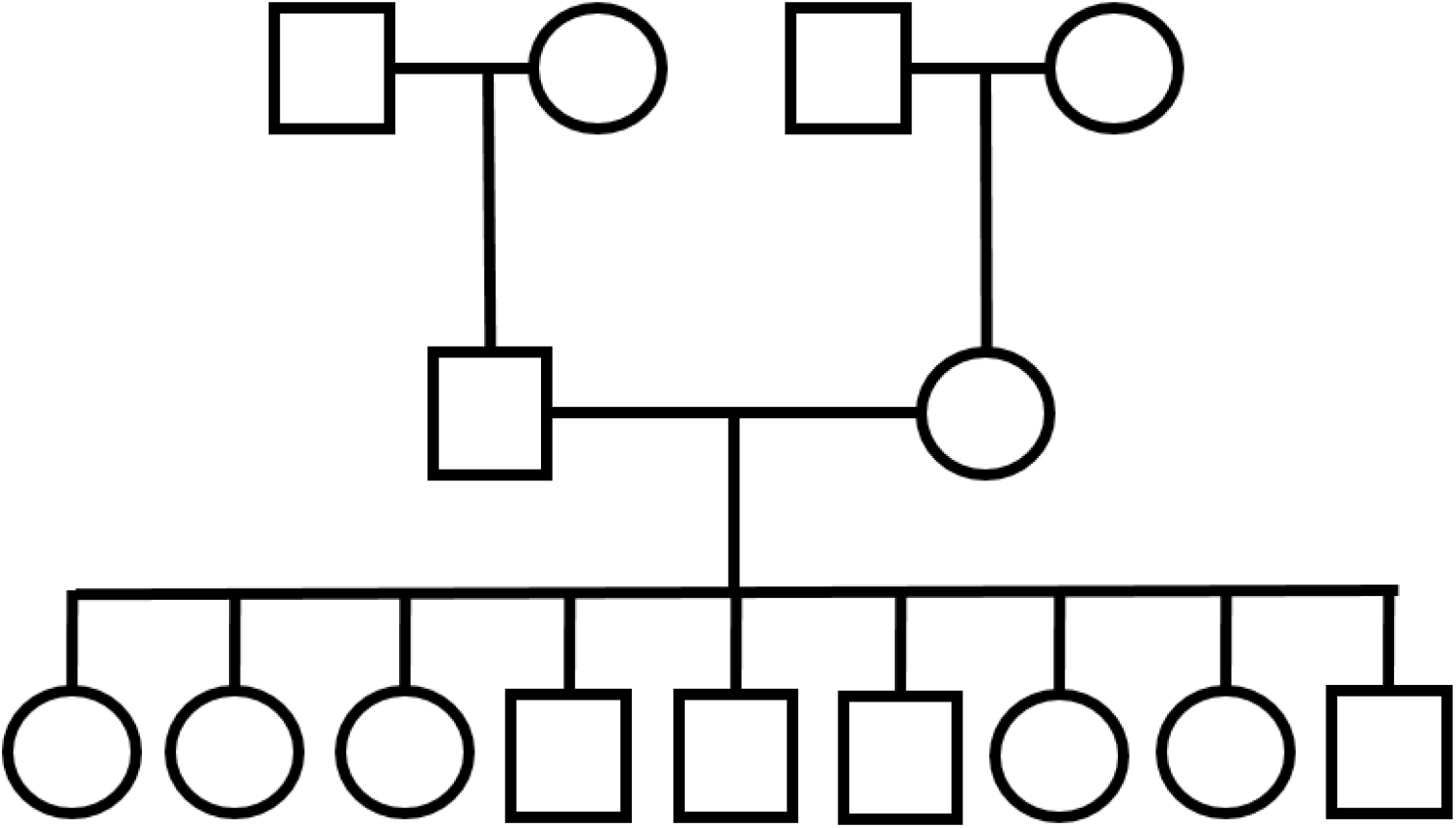
Utah Genome Reference Project representative pedigree Caption: This pedigree illustrates the family structure of participants. Families were centered on the middle generation, both of whose parents and children were enrolled. For the purpose of genetic risk score estimation, any participant with available parental DNA were analyzed as offspring, including those of the middle generation.

The GRSs for each trait were calculated for all participants with available genotype data. Calculated GRSs for offspring served as a reference against which the parentally-derived estimated GRSs would be assessed. “Raw” GRSs considered each SNP as contributing equally to the trait, with a score of +1 for the alternate allele with a positive effect and -1 for the alternate allele with a negative effect. Weighted GRSs incorporated each SNP’s beta coefficient, which weights each SNP according to the magnitude of its effect. Estimated weighted and raw offspring GRSs were calculated from parental SNP data using a weighted-averages approach and by computing the simple average of the two parental GRSs. Regarding terminology, “weighted GRS” refers to a GRS wherein each SNP’s score is multiplied by a coefficient that corresponds to its effect size for the given trait (its “weight”), whereas the “weighted-average method” refers to the method of estimating offspring GRS from the weighted average of all possible scores transmitted from parents at a given SNP, which is described in detail below.

Both weighted and raw estimated GRSs were calculated by the weighted-averages method as follows. From one set of parents, all possible transmitted allele combinations at a given SNP were used to calculate all possible transmitted scores at that SNP. Each transmitted alternate allele confers a score of 1 or -1 depending on the direction of effect, and the reference allele a score of 0. Each possible transmitted score was multiplied by the probability of its transmission, and the products were summed to generate the estimated GRS for the SNP. Estimated scores for SNPs were summed for each trait’s estimated GRS. The For example, at a SNP with a positive effect on a trait, a heterozygous paternal genotype paired with the same maternal genotype has a 25% chance of transmitting both alternate alleles (score = +2), a 50% chance of one alternate and one reference allele (score = +1), and a 25% chance of two reference alleles (score = 0), as illustrated with a simple Punnett square (Figure 2). This is based on a simple assumption of genetic recombination and passing a single allele to each offspring. The estimated GRS at this SNP for offspring of this parental pair would therefore be expressed as 0.25(2) + 0.5(1) + 0.25(0). The GRS for a given trait is then computed as the sum of the estimated scores from each SNP. This method was used to calculate both raw and weighted GRSs for each offspring participant for each of the preceding traits. Parental genotypes were also used to compute the minimum and maximum GRSs that could be transmitted to offspring.

**Figure 2.**
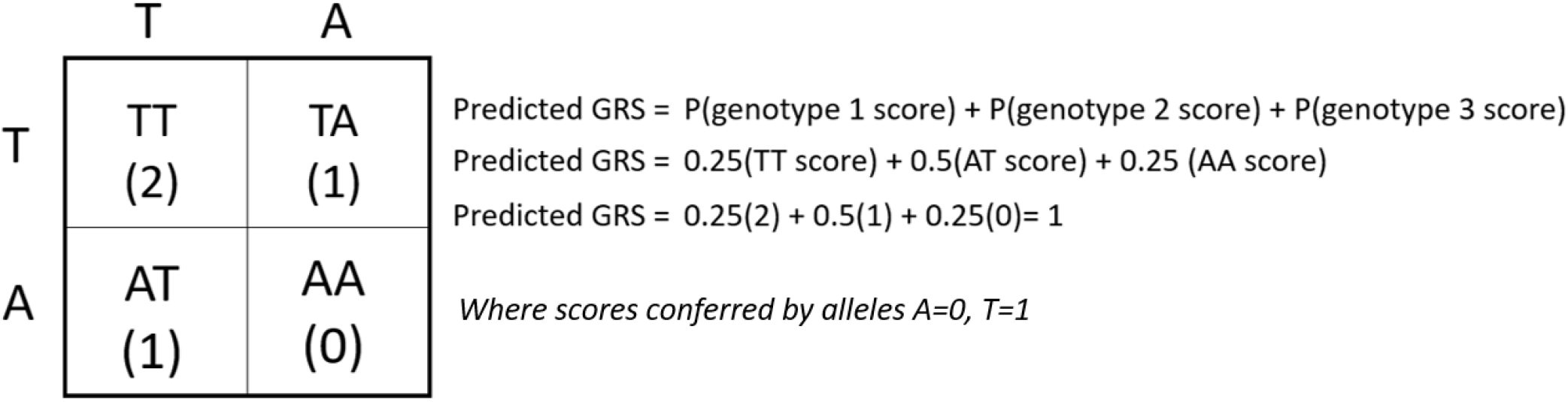
Illustration of offspring GRS estimation for one SNP Caption: Punnett square and offspring estimated GRS calculation at a single SNP site from two heterozygous parents, where each possible score is weighted according to the probability of its transmission. GRS, genetic risk score; SNP, single nucleotide polymorphism.

The primary outcome was the percent error of the estimated GRS for each trait. Percent error was defined as the discrepancy between actual and estimated offspring GRSs, divided by the range of possible inherited GRSs from each offspring’s parents for that trait. Predictions yielding mean percent errors < 10% were considered accurate. Accuracy was assessed at the level of the cohort by comparison of mean estimated vs mean actual GRSs using the t-test. Secondary analyses included proportion of participants with percent errors less than 10%, 15%, and 20%, comparison of percent errors between weighted average and simple average approaches, accuracy of estimated offspring GRSs using DNA from only one parent and the cohort average for the missing parent, and accuracy of GRS based in association with fetal sex and number of SNPs used to compute the GRS. All analyses were carried out for both raw and weighted GRSs. GRSs were computed using the list of SNPs and beta coefficients reported by Chen et al^10^. Finally, we used linear mixed modeling at the level of the family to assess for associations between GRS estimation accuracy and offspring sex and birth order. The threshold for significance was a two-sided p value of 0.05. Analyses were carried out using Stata^11^.

## Results

A total of 43 families (N=454 offspring) that had both parental and offspring data available were included in this analysis. Families had an average of 8.9 ± 1.8 offspring each. GRSs were based on the following number of SNPs: height: 2,130; BP: 831; BMI: 628; T2D: 306; BW: 86; FPG: 22. Characteristics of the study participants are summarized in Table 1.

**Table 1.** Cohort characteristics

|  |  |
| --- | --- |
|  | N=454 offspring |
| Female sex, n(%) | 232 (50.9) |
| Offspring per family, mean SD | 8.9 ± 1.8 |
| Birth year (mean, SD) | 1966 ± 13.4 |
| Height |  |
| SNPs | 2,130 |
| Raw GRS, mean (min, max) | 30.0 (-62, 108) |
| Wtd GRS, mean (min, max) | -0.16 (-1.9, 1.15) |
| Blood pressure |  |
| SNPs | 831 |
| Raw GRS, mean (min, max) | -10.7 (-70, 39) |
| Wtd GRS mean (min, max) | 1.16 (-8.4, 9.05) |
| BMI |  |
| SNPs | 628 |
| Raw GRS, mean (min, max) | -25.5 (-71, 29) |
| Wtd GRS, mean (min, max) | -0.04 (-0.78, 0.71) |
| Type 2 diabetes mellitus |  |
| SNPs | 306 |
| Raw GRS, mean (min, max) | -14.1 (-41, 15) |
| Wtd GRS, mean (min, max) | -0.29 (-0.93, 0.37) |
| Birth weight |  |
| SNPs | 86 |
| Raw GRS, mean (min, max) | 10.4 (-6, 24) |
| Wtd GRS, mean (min, max) | 0.17 (-0.22, 0.49) |
| Fasting plasma glucose |  |
| SNPs | 22 |
| Raw GRS, mean (min, max) | 2.3 (-8, 12) |
| Wtd GRS, mean (min, max) | 0.13 (-0.22, 0.45) |

Abbreviations: SNP, single nucleotide polymorphisms; GRS, genetic risk score

Mean percent errors for both weighted and raw estimated offspring GRSs were less than 10% for all traits but FPG (Table 2). Mean percent errors were smallest for the traits of height (1.1% ± 0.8) and blood pressure (1.6% ± 1.2) and largest for the trait of plasma fasting glucose (10.5% ± 8.1%, Table A).

**Table 2.** Percent error of both raw and weighted offspring estimated GRSs.

| Trait | Raw Estimated<br>GRS percent error | Weighted Estimated<br>GRS percent error |
| --- | --- | --- |
| BW | 5.1 ±4.0 | 5.8 ±4.2 |
| Height | 1.1±0.8 | 1.3 ±1.0 |
| BMI | 1.8±1.4 | 2.1 ±1.5 |
| T2D | 2.7±2.1 | 3.1 ±2.4 |
| FPG | 10.5±8.1 | 12.1 ±9.2 |
| BP | 1.6±1.2 | 1.9 ±1.4 |

Abbreviations: BW, birth weight; BMI, body mass index; T2D, type 2 diabetes; FPG, fasting plasma glucose; BP, blood pressure
Data are expressed as mean ± standard deviation.

When plotted against the number of SNPs within each trait, percent error decreased exponentially with increasing SNPs per trait with diminishing improvement in percent error above 500 SNPs (Figure 3). Estimated raw and weighted GRSs were not statistically different from actual GRSs for all traits evaluated (Table 3). Estimation of offspring GRSs for height, BMI, and BP were accurate in 100% of offspring, while BW GRS estimation was accurate in 88% (95% CI, 85-91 and FPG in 53% (95% CI 48-58). As expected, the proportion of offspring with accurately estimated GRSs for BW and FPG increased progressively at accuracy thresholds of 15% and 20%. Proportions of offspring with accurately estimated GRSs are summarized in Table 4. Estimating GRS using the simple average of the maternal and paternal GRS for a given trait yielded exactly the same values and distributions as the weighted average approach (Table 5).

**Figure 3.**
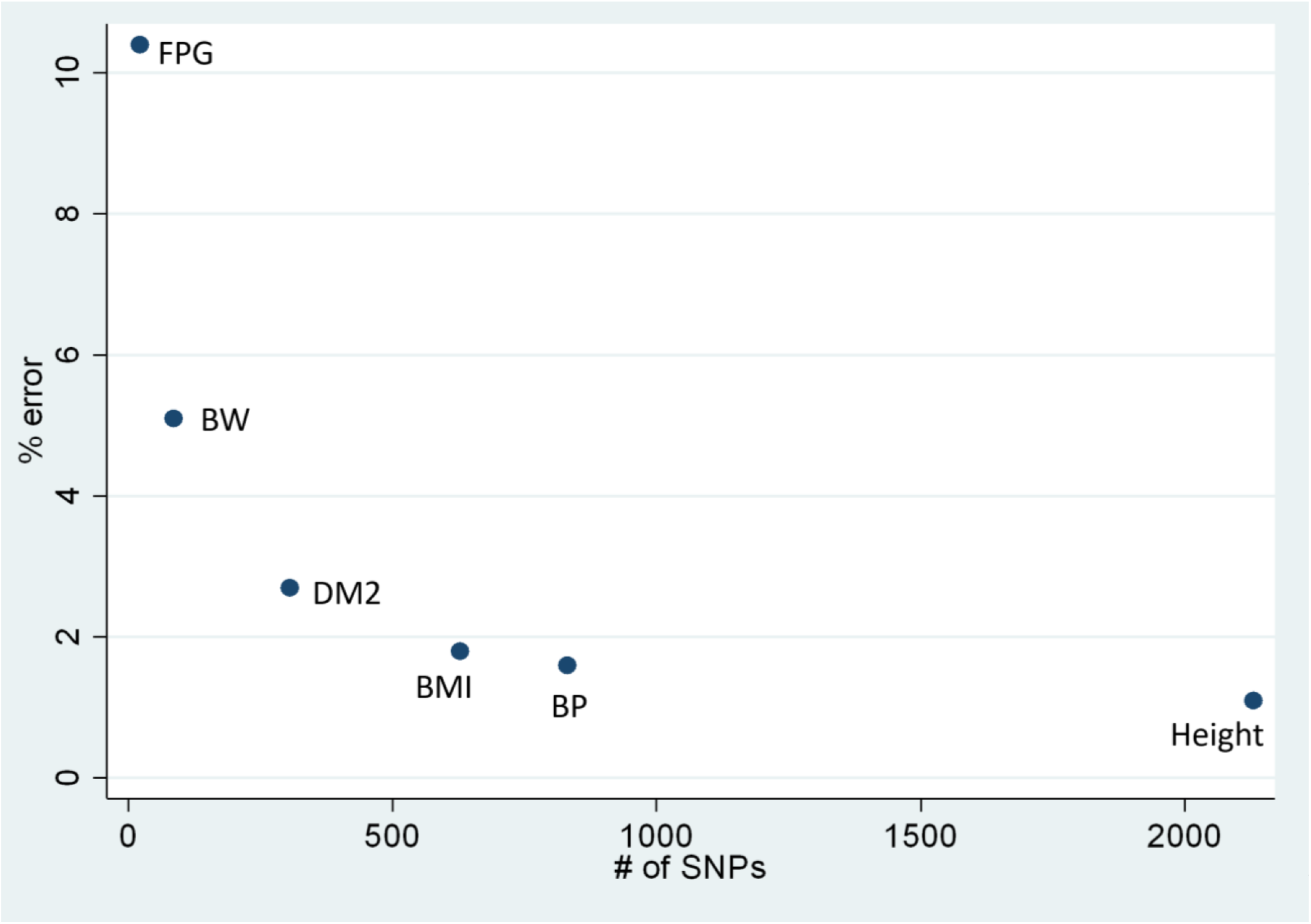
Percent error as a function of number of SNPs in each trait’s GRS Caption: Scatter plot demonstrating mean percent error between estimated versus actual genetic risk scores as a function of the total number of SNPs available for genetic risk score calculations. Abbreviation: SNP, single nucleotide polymorphism; GRS, genetic risk score; FPG, fasting plasma glucose; BW, birth weight; DM2, type 2 diabetes mellitus; BMI, body mass index; BP, blood pressure.

**Table 3.** Comparison of actual versus estimated offspring raw GRSs

| Trait | Actual GRS (CI) | Estimated GRS (CI) | P value* |
| --- | --- | --- | --- |
| BW | 10.4 (10.0-10.9) | 10.2 (9.9-10.5) | 0.5 |
| Height | 30.0 (27.5-32.4) | 29.1 (27.6-30.7) | 0.6 |
| BMI | -25.5 (-27.1- -23.9) | -25.9 (-27.0- -24.8) | 0.7 |
| T2D | -14.1 (-14.9- -13.2) | -14.2 (-14.8- -13.7) | 0.8 |
| PFG | 2.3 (2.0-2.6) | 2.4. (2.2-2.6) | 0.6 |
| BP | -10.7 (-12.4- -9.0) | -10.1 (-11.2- -8.9) | 0.6 |

Abbreviations: BW, birth weight; BMI, body mass index; T2D, type 2 diabetes; FPG, fasting plasma glucose; BP, blood pressure
Data are expressed as mean (95% confidence interval).

**Table 4.** Proportions of participants with GRSs estimated ac varying thresholds of accuracy.

| Trait | Accurate to within 10% | Accurate to within 15% | Accurate to within 20% |
| --- | --- | --- | --- |
| BW | .88 (0.85-0.91) | 0.97 (0.95-0.98) | 0.99 (0.98-0.99) |
| Height | 1.0 (--) | 1.0 (--) | 1.0 (--) |
| BMI | 1.0 (--) | 1.0 (--) | 1.0 (--) |
| T2D | 0.99 (0.98-0.99) | 1.0 (--) | 1.0 (--) |
| PFG | 0.53 (0.48-0.58) | 0.75 (0.71-0.79) | 0.86 (0.83-0.89) |
| BP | 1.0 (--) | 1.0 (--) | 1.0 (--) |

Abbreviations: BW, birth weight; BMI, body mass index; T2D, type 2 diabetes; FPG, fasting plasma glucose; BP, blood pressure
Data are expressed as: proportion (95% confidence interval)

**Table 5.** Comparison of estimated offspring GRSs using weighted average versus simple average methods.

| Trait | Pred GRS weighted avg | Pred GRS simple avg | P value |
| --- | --- | --- | --- |
| BW | 10.2 ± 3.3 | 10.2 ± 3.3 | -- |
| Height | 29.4 ± 17.0 | 29.4 ± 17.0 | -- |
| BMI | -25.9 ± 12.5 | -25.9 ± 12.5 | -- |
| T2D | -14.2 ± 6.1 | -14.2 ± 6.1 | -- |
| PFG | 2.4. ± 2.2 | 2.3 ± 2.2 | -- |
| BP | -10.2 ± 12.8 | -10.2 ± 12.8 | -- |

Abbreviations: BW, birth weight; BMI, body mass index; T2D, type 2 diabetes; FPG, fasting plasma glucose; BP, blood pressure
Data are expressed as mean ± standard deviation

When offspring GRSs were estimated with a simulated missing parent by substituting the cohort mean for males for a missing father and female mean GRS for missing mothers, there were still no significant differences between actual and estimated GRSs (Appendix). Percent errors for estimated GRSs were slightly higher when paternal or maternal DNA were not available, but the differences were small; mean differences were <1% for all traits but FPG (Table 6), such that proportions of offspring with accurately estimated GRSs were similar to when both parental GRSs were used (Appendix). Percent errors were not consistently better when paternal DNA was missing when compared to maternal DNA (Table 7). Using linear mixed effects modeling, neither birth order nor offspring sex were associated with percent error of estimated GRSs (data not shown).

**Table 6.** Comparison of estimated offspring raw GRSs using DNA from both parents versus with one parent missing

| Trait | % error, both<br>parents available<br>Mean, SD | % error, no paternal<br>DNA available<br>Mean, SD | Mean difference | P value |
| --- | --- | --- | --- | --- |
| Height | 1.1 ±0.9 | 1.4 ±1.0 | 0.3 | <0.001 |
| BP | 1.6 ±1.2 | 1.9 ±1.5 | 0.3 | <0.001 |
| BMI | 1.8 ±1.4 | 2.4 ±1.8 | 0.6 | <0.001 |
| T2D | 2.7 ±2.1 | 3.3 ±2.5 | 0.6 | <0.001 |
| BW | 5.1 ±4.0 | 6.0 ±4.4 | 0.8 | 0.003 |
| PFG | 10.5 ±8.1 | 13.7 ±10.7 | 3.2 | <0.001 |
| Trait | % error, both<br>parents available<br>Mean, SD | % error, no maternal<br>DNA available<br>Mean, SD | Mean difference | P value |
| Height | 1.1 ±1.0 | 1.2 ±1.0 | 0.1 | 0.03 |
| BP | 1.6 ±1.2 | 2.1 ±1.7 | 0.5 | <0.001 |
| BMI | 1.8 ±1.4 | 2.4 ±1.8 | 0.6 | <0.001 |
| T2D | 2.7 ±2.1 | 3.1 ±2.4 | 0.3 | 0.05 |
| BW | 5.1 ±4.0 | 5.9 ±4.4 | 0.8 | 0.04 |
| PFG | 10.5 ±8.1 | 13.0 ±10.0 | 2.5 | <0.001 |

Abbreviations: BW, birth weight; BMI, body mass index; T2D, type 2 diabetes; FPG, fasting plasma glucose; BP, blood pressure
Data are expressed as mean ± standard deviation

**Table 7.** Comparison of estimated raw GRS percent errors with missing paternal versus missing maternal DNA

| Trait | Percent error with missing paternal DNA (%) | Percent error with missing maternal DNA (%) | Mean difference (%) | P value |
| --- | --- | --- | --- | --- |
| Height | 1.4 ± 1.0 | 1.2 ± 1.0 | -0.1 | 0.04 |
| BP | 2.1 ± 1.5 | 2.0 ± 1.7 | 0.18 | 0.07 |
| BMI | 2.4 ± 1.8 | 2.4 ± 1.8 | -0.01 | 0.92 |
| T2D | 3.3 ± 2.5 | 3.0 ± 2.4 | -0.31 | 0.06 |
| BW | 5.9 ± 4.4 | 5.9 ± 4.4 | -0.01 | 0.98 |
| PFG | 13.0 ± 10.0 | 13.7 ± 10.6 | -0.7 | 0.3 |

Abbreviations: BW, birth weight; BMI, body mass index; T2D, type 2 diabetes; FPG, fasting plasma glucose; BP, blood pressure
Data are expressed as mean ± standard deviation

## Discussion

Using genomic data from a large cohort of multigenerational families, we demonstrated that offspring genetic risk scores can be estimated using parental SNP array data. Estimated GRSs had mean percent errors less than 10% for all traits but FPG, with accuracy improving with increasing numbers of SNPs within each trait. We showed that estimated GRSs can be computed from taking a simple average of the two parents’ GRSs, making it easy to carry out in contexts without dedicated computational resources or tools. When only one parent’s DNA is available, estimation was accuracy was statistically worse, though the difference in mean percent error was <1% for all traits but FPG. While our analysis only evaluated GRSs for 6 traits, they are likely applicable to other traits for which GRSs derived from primarily autosomal SNPs.

Our findings are relevant to the changing landscape of prenatal diagnosis modalities. While genetic risk scores are not routinely used in clinical obstetrics, they have potential to be informative for clinical management. There are a variety of multifactorial outcomes for which risk stratification is based on population-level data but could be better individualized by knowledge of the fetal GRS for that trait. Perhaps the best example is fetal growth potential, which is currently assessed based on population norms but is presumed to be dependent on genetic factors. Fetal growth restriction is conceptually defined as a fetus’s inability to meet its growth potential. In practice, the individual fetus’s growth potential is unknown. Accordingly, a fetus labeled “small” by population standards may actually be healthy, while another large fetus who has failed to achieve their true growth potential may be labeled “normal” but be at increased risk of adverse outcomes. Other such opportunities are easy to conceive, such as responsiveness to antenatal corticosteroids or risk stratification for neonatal complications of late preterm delivery. Knowledge of fetal genetic risk scores may offer promise to individualize such assessments and has the potential to improve care. While these opportunities are currently hypothetical, we describe a means by which to investigate them without the significant obstacle of directly interrogating the fetal genome to compute GRSs.

The recent emergence of commercial GRS use for human embryo selection following *in vitro* fertilization is problematic for multiple reasons, not the least of which are ethical concerns related to selection based on inadequately characterized probabilities of more or less desirable adult traits(4). Although prenatal use of GRSs is not automatically protected from misuse, its prospect as a means to individualize care is legitimate insofar as it is appropriately investigated and validated prior to clinical application.

Our study had limitations. The primary limitation is the fact that findings from this cohort, which was recruited from a racially homogeneous group, may not be generalizable to more diverse populations. However, our analysis does not depend on the frequency of specific variants or their associations with traits, which are known to vary across racial groups. Rather, our analysis is merely a demonstration that principles of mathematical probability can be used to estimate offspring GRSs. It still is necessary to validate and refine this approach using larger and more diverse cohorts. Our study also had strengths. The large family size and systematic sampling of DNA from all available children allowed for more robust testing for association between estimation accuracy with offspring sex and birth order using within-family analysis.

In summary, we demonstrated that offspring GRSs can be estimated using a simple average of two biological parents, and that estimation suffers only marginally when DNA is only available from one parent. The specific traits used in this analysis are less important than the method itself since the pattern of accuracy follows that which would be predicted by the central limit theorem. Validation of this approach is necessary for traits or conditions with SNPs on sex chromosomes and in more diverse cohorts before broader testing and application.

## Data Availability

Whole genome sequencing data for UGRP cohort participants are available in CRAM and VCF format via controlled access through the University of Utah Center for Genomic Medicine. Overview of available data and the application process are available at https://uofuhealth.utah.edu/center-genomic-medicine/research/ceph-resources.php.

## Acknowledgements

We express our thanks to the Utah CEPH consortium participants, as well as to Ray White, Jean-Marc Lalouel, Stephen M. Prescott, and Mark Leppert, who were instrumental in the ascertainment of the CEPH/Utah pedigrees.

---

**Table S1.** Comparison of actual GRS with predicted GRS with missing paternal DNA

| Trait | Actual GRS (CI) | Predicted GRS NO DAD (CI) | P value |
| --- | --- | --- | --- |
| BW | 10.4 (10.0-10.9) | 10.2(10.0-10.4) | 0.4 |
| Height | 30.0 (27.5-32.5) | 29.3 (28.2-30.5) | 0.6 |
| BMI | -25.5 (-27.1- -23.9) | -25.9 (-26.8- -25.1) | 0.7 |
| T2D | -14.1 (-15.0- -13.2) | -14.2 (-14.6- -13.8) | 0.8 |
| PFG | 2.3 (2.0-2.6) | 2.4. (2.2-2.5) | 0.6 |
| BP | -10.7 (-12.4- -9.0) | -10.1 (-11.2—8.9) | 0.6 |

**Table S2:** comparison of actual vs estimated GRS with missing maternal DNA

| Trait | Actual GRS (CI) | Predicted GRS NO MOM (CI) | P value |
| --- | --- | --- | --- |
| BW | 10.5 (10.0-10.9) | 10.2 (10.0-10.4) | 0.3 |
| Height | 30.0 (27.5-32.5) | 29.3 (28.1-30.6) | 0.6 |
| BMI | -25.6 (-27.2- -24.0) | -25.9 (-26.7- -25.1) | 0.7 |
| T2D | -14.1 (-14.9- -13.2) | -14.2 (-14.7- -13.8) | 0.7 |
| PFG | 2.3 (2.0-2.6) | 2.4. (2.2-2.5) | 0.6 |
| BP | -10.9 (-12.7- -9.2) | -10.1 (-10.9- -9.4) | 0.4 |

**Table S3:** Accuracy when no paternal info is available:

| Trait | Accurate to within 10% (95% CI) | Accurate to within 15% (95% CI) | Accurate to within 20% (95% CI) |
| --- | --- | --- | --- |
| BW | .84 (0.81-0.87) | 0.95 (0.93-0.97) | 1.0 (0.98-1.0) |
| Height | 1.0 (--) | 1.0 (--) | 1.0 (--) |
| BMI | 1.0 (--) | 1.0 (--) | 1.0 (--) |
| T2D | 0.99 (0.98-0.99) | 1.0 (--) | 1.0 (--) |
| PFG | 0.45 (0.41-0.50) | 0.62 (0.58-0.67) | 0.76 (0.72-0.80) |
| BP | 1.0 (--) | 1.0 (--) | 1.0 (--) |

**Table S4:** Accuracy when no maternal info is available:

| Trait | Accurate to within 10% (95% CI) | Accurate to within 15% (95% CI) | Accurate to within 20% (95% CI) |
| --- | --- | --- | --- |
| BW | .79 (0.78-0.83) | 0.96 (0.94-0.98) | 0.99 (0.98-1.0) |
| Height | 1.0 (--) | 1.0 (--) | 1.0 (--) |
| BMI | 1.0 (--) | 1.0 (--) | 1.0 (--) |
| T2D | 0.99 (0.97-1.0) | 1.0 (--) | 1.0 (--) |
| PFG | 0.46 (0.42-0.51) | 0.62 (0.57-0.66) | 0.79 (0.75-0.83) |
| BP | 1.0 (--) | 1.0 (--) | 1.0 (--) |

